# Detecting measles virus in hospital air during a local outbreak

**DOI:** 10.64898/2026.09.25.26363929

**Authors:** Hannah J. Barbian, Michelle Funk, Stephanie R. Black, Mary K. Hayden, Shane Zelencik, Brian F. Borah, David Zhang, Allison H. Bartlett, Larry K. Kociolek, Ayelet Rosenthal, Nabgha Farhat, V. Eloesa McSorley, Alyse Kittner

**Affiliations:** Division of Infectious Diseases, Department of Internal Medicine, Rush University Medical Center, Chicago, Illinois, USA; Disease Control and Emergency Preparedness Bureau, Chicago Department of Public Health, Chicago, Illinois, USA; Department of Pediatrics, Section of Pediatric Infectious Diseases, Comer Children’s Hospital, University of Chicago, Chicago, Illinois, USA; Division of Pediatric Infectious Diseases, Ann & Robert H. Lurie Children’s Hospital of Chicago; Department of Pediatrics, Northwestern University Feinberg School of Medicine, Chicago, Illinois, USA

## Abstract

In 2024, Chicago experienced a measles virus outbreak. Air samples collected as part of routine respiratory virus surveillance from three hospitals that received measles patients were retrospectively analyzed. Measles virus was detected in air samples from one emergency department across five weeks when measles patients were present and another when no known cases were present. Two of three viral sequences derived from air samples were identical to measles outbreak patients. The source of one diverging sequence could not be identified and may represent a case not identified by traditional surveillance methods. Air sampling may supplement measles investigation, surveillance and response.

## BACKGROUND

Measles virus is a highly contagious pathogen that is transmitted through air. Measles outbreaks are increasing in the U.S.; additional tools to aid measles surveillance could help contain outbreaks. Because measles’ route of transmission is airborne, indoor air sampling may be a useful tool for measles early detection, infection prevention, outbreak investigation and response. Preliminary work shows that measles virus RNA is detectable in hospital rooms containing infected patients in as little as 30 minutes of air sample collection, but it is unknown whether air sampling can detect measles cases in healthcare or public settings where cases are transiently present.^1,2^

In 2024, Chicago experienced a measles outbreak associated with a large shelter for newly arrived migrants; 66 cases among Illinois residents were identified from February through April.^3^ In total, 52 case-patients were hospitalized across six hospitals; sufficient isolation space was unavailable in the congregate shelter, so many measles-positive shelter residents were hospitalized strictly for isolation and without a medical indication.^3,4^

Since 2023, Chicago Department of Public Health has managed an indoor air surveillance program to monitor for seasonal respiratory pathogens.^5,6^ As part of this program, air samplers placed in hospital emergency departments (EDs) collected weekly samples; three of these hospitals admitted measles patients during the 2024 outbreak. Here, we performed a retrospective analysis testing these air samples for the presence of measles virus to inform whether indoor air sampling may be useful as a surveillance or investigative tool for measles virus exposures.

## METHODS

### Air sample collection

Air surveillance was performed as part of public health surveillance activities by the Chicago Department of Public Health. This study was determined to be exempt from IRB review under federal regulation 45 CFR 46.102(l)(2). Four AerosolSense samplers (ThermoFisher) were located at three emergency departments (ED1 - ED3). The ED1 sampler was adjacent to a triage station near patient rooms. The ED2 sampler was in the waiting room near patient seating. ED3 had two samplers, one in a triage station near patient rooms and another near the waiting room entrance. All samplers were placed at table height. AerosolSense cartridges were inserted and collected at an airflow rate of 200 L/min for 7 continuous days. On day 7, new cartridges were immediately inserted for the next sampling week. Cartridges were transported via local courier to the Regional Innovative Public Health Laboratory, a contracted laboratory network partner of Chicago Department of Public Health at Rush University Medical Center, and stored at 4°C.

### Molecular detection of measles virus

AerosolSense cartridges contain two membranes; one was immediately processed as part of routine respiratory virus surveillance and residual extract was stored at -80°C.^5^ The second membrane was stored at -80°C. Total nucleic acids were extracted from air cartridges membranes as previously described.^7^ Quantitative PCR (qPCR) assays were used for detection of measles virus nucleoprotein gene^8^ and measles virus vaccine strain.^9^ TaqPath 1-Step Multiplex Master Mix (ThermoFisher) and 5 µl air sample extract was used for each reaction in duplicate. Assay limit of detection was determined by spiking a limiting dilution of synthetic measles DNA into negative control air samples collected in a storage room. The mean Ct value of the minimal dilution where 90% of 10 replicates yielded detectable amplification was used as a cutoff; Ct values ≤35.7 were considered “positive”, Ct values >35.7 were considered “negative”. Total nucleic acid extraction and measles qPCR was repeated on the second cartridge membrane for any sample with detectable Ct values (<40).

### N450 sequencing and analysis

For increased sensitivity from indoor air samples, a nested PCR was adapted from an N450 international benchmark standard sequencing protocol.^10^ Reverse transcription was performed using the outer reverse primer and Superscript IV Reverse Transcriptase (ThermoFisher). The first round of PCR was performed as described^10^ using repliQa HiFi ToughMix (Quantabio) and 1 μL cDNA; the second round of PCR was performed using 1 μL of the first-round product, forward primer 5-GGAGGTCAGCTGGRAAGGTCAG-3, reverse primer 5-GGGTAGGYRGATGTTGTTCTGG-3, and 25 cycles of PCR with an annealing temperature of 52°C. Adapter sequences were added to inner primers for direct amplicon sequencing.^11^ Sequencing was performed on Illumina MiSeq i100 2×300; sequencing depth was >30,000 for all samples. Primer-trimmed reads were assembled to measles reference sequence (Genbank accession K01711.1). Genbank accessions for N450 sequences derived from air samples are QB065539 - QB065541.

Sequences from clinical samples were acquired from NCBI Virus (https://www.ncbi.nlm.nih.gov/labs/virus/vssi/). Phylogenies were produced using IQTree.

## RESULTS

Air samples collected between January 26, 2024 (one month before the first reported measles case in Illinois) and May 27, 2024 (one month following the last reported outbreak case) were included in this study. A total of 73 air samples from four air samplers in three EDs were analyzed. There were 29 patients with measles admitted to these three facilities during this time; the ED served as the point of entry for 27 patients via emergency medical services (n=26) or waiting room (n=1); 2 patients admitted at the hospital associated with ED3 did not enter the ED.

Measles RNA was detected in 5 consecutive air samples from ED1, with sampling dates spanning March 20, 2024 to April 23, 2024 (Figure 1, Supplemental Table 1). This corresponded with peak measles virus transmission in Chicago, as well as 11 measles patient hospitalizations at the ED1 facility (Figure 1). One air sample from ED2 was positive with the sampling date beginning six days after the rash onset date of the last reported measles case in an Illinois resident and when no measles patients were hospitalized at the facility (Figure 1). All air samples collected from two air samplers in ED3 were negative (Figure 1). All air samples were negative for measles vaccine strain, indicating wild type measles detection in ED1 and ED2 (Supplementary Table 1).

**Figure 1.**
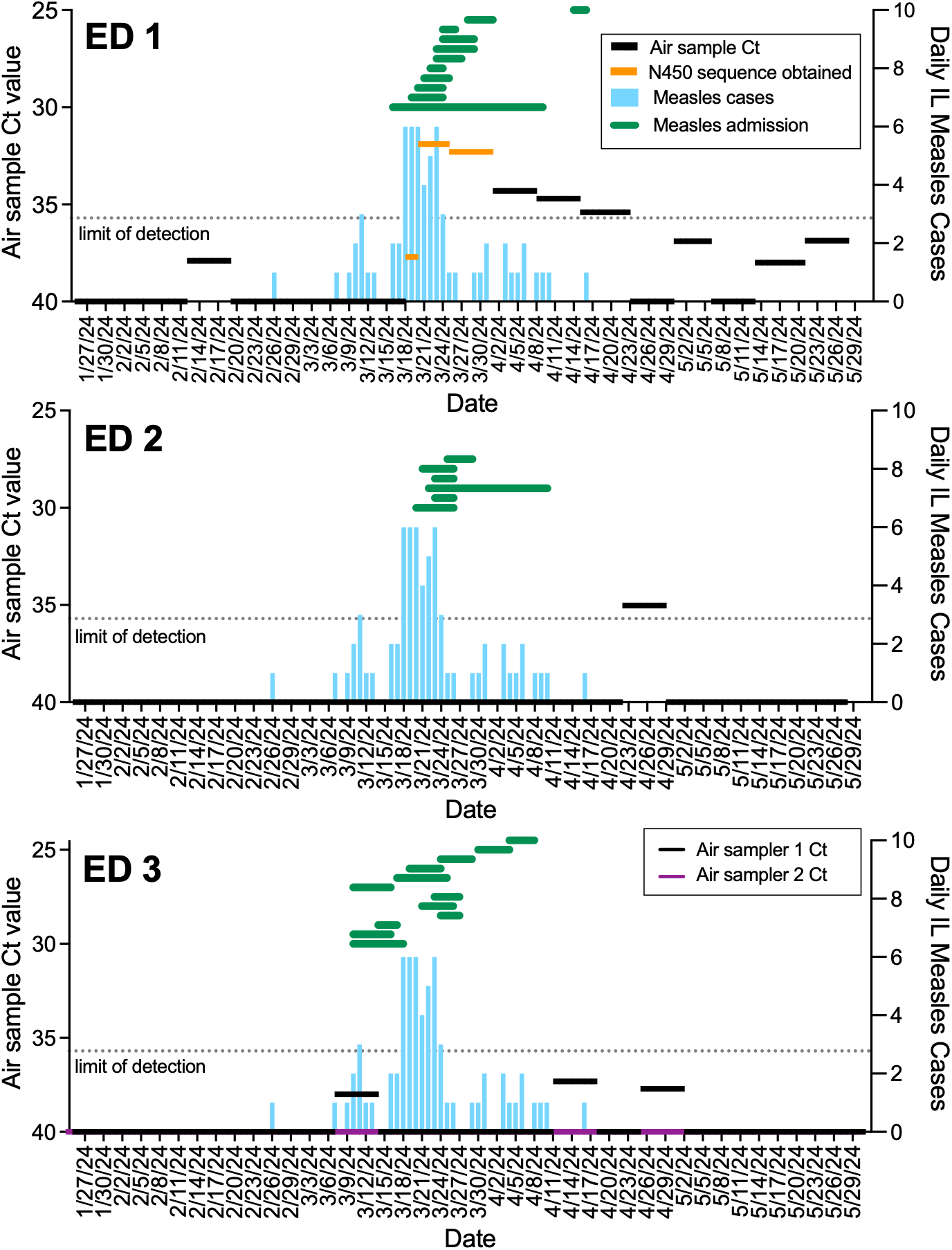
Detection of measles virus in hospital air. Air sample Ct values for measles virus at three emergency departments are shown in black horizontal lines (ED 1, ED 2, and ED 3 panels, left y axis). Most air samples were collected for 7 continuous days; the timeframe of sampling is indicated by the corresponding x axis dates. ED3 (bottom panel) had two air samplers, results of the second sampler are shown in purple. Air samples where measles N450 were able to be sequenced are shown in orange. Daily Illinois resident measles incidence is shown in blue columns, by rash onset date (right y axis). Measles admissions in the same hospital as the corresponding ED are overlaid in green horizontal lines, with start and end dates indicating case admission and discharge. Dashed horizontal line indicates the limit of detection Ct value cutoff.

Measles N450 sequences, the international standard for measles virus genotyping, were obtained from three air samples from ED1, including two with the lowest measles Ct values and one with a Ct value above the detection cutoff (Figure 1). Air sample N450 sequences were genotype D8, the measles virus genotype present in the 2024 Chicago outbreak.^3^ Compared to N450 sequences collected from U.S. clinical cases in 2024, air sample sequences clustered with Illinois outbreak-associated sequences, with 2 air samples being identical to contemporaneous Illinois clinical sequences, and 1 air sample being 2 single nucleotide polymorphisms diverged (Figure 2). N450 sequences could not be obtained from other air samples with detectable measles virus (Supplemental Table 1).

**Figure 2.**
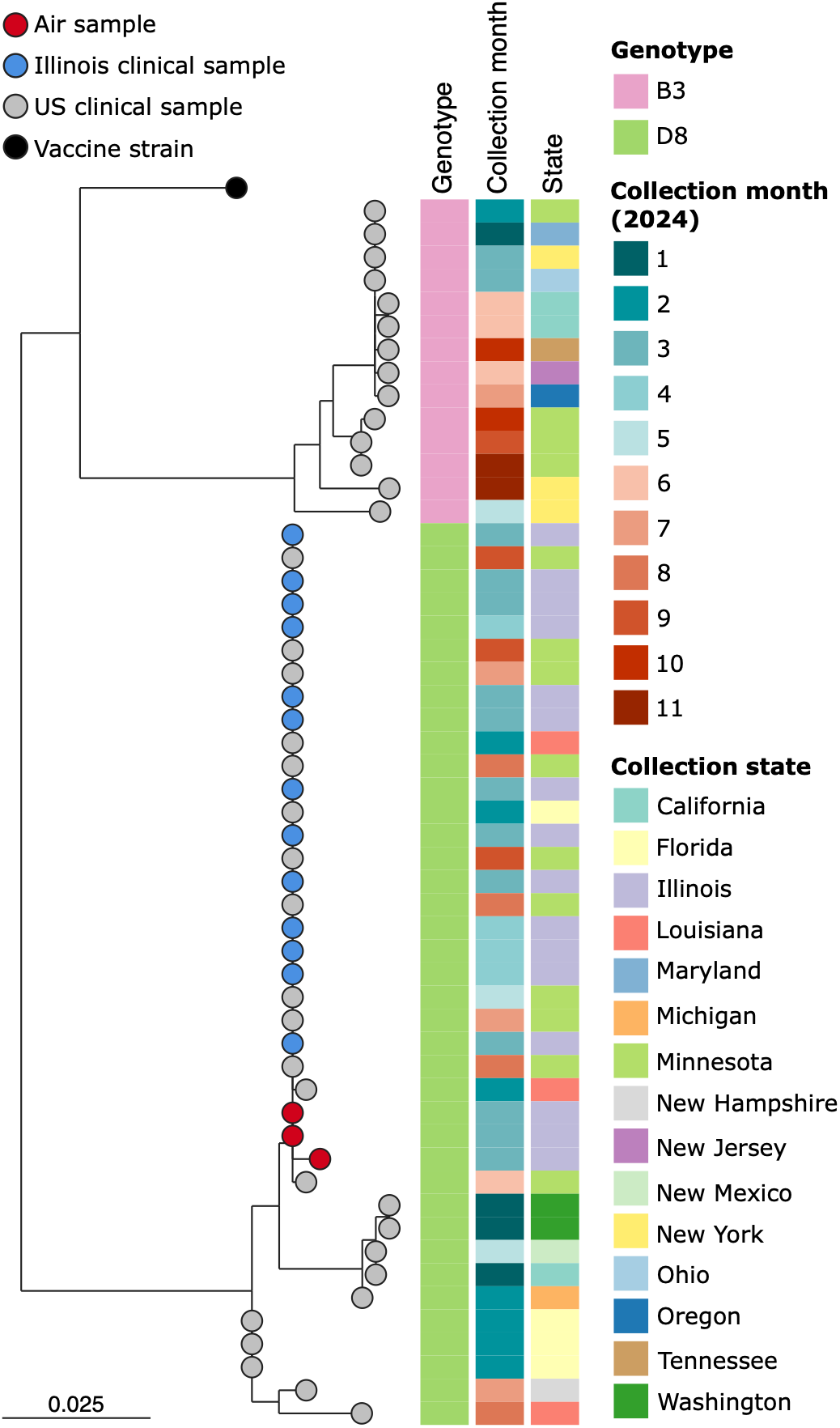
Phylogeny of measles virus collected from indoor air with concurrent clinical samples. Maximum-likelihood phylogeny of measles N450 region for air samples collected in this study (red nodes) and randomly sampled sequences from U.S. 2024 clinical samples (blue nodes = Illinois, grey nodes = other US states). Metadata to the right of tree nodes indicate sequence genotype, month of specimen collection, and US state of specimen collection. Scale indicates substitutions per site.

## DISCUSSION

We detected measles virus nucleic acids in the air of two of the three hospital EDs with air samplers that received measles patients during a local outbreak. In one hospital (ED1), detections were continuous across five consecutive weeks. Due to the large volume of measles patients seen by this hospital, some measles patients were housed in negative pressure but non-airborne infection isolation rooms within the ED. The air sampler in this hospital was placed ∼30 feet from one such room where at least one measles patient was present across sampling weeks when measles virus was detected. Notably, this room did not contain an isolation anteroom, and while measles patients did not leave the room, staff and family members entered and exited as needed. Air escape during entry/exit may explain measles virus detection in the adjacent hallway. Measles patients were transported in isolation tents in this facility; however, facility staff noted at least one instance where an infant was transported from the ambulance bay to a triage station adjacent to the air sampler without an isolation tent. This may also have contributed to measles detection in the air. Another ED (ED3) had an air sampler located in a similar position as ED1, *i*.*e*. near a triage station adjacent to treatment rooms, however, this ED utilized only airborne infection isolation rooms for measles positive patients and the air sampler was not located nearby any of these rooms. There were no measles detections in this ED.

Measles virus was detected in one air sample from another ED (ED2) when no measles patients were known to be present in the facility and after the last reported case in an Illinois resident. The nucleic acid level was near the limit of detection and could have represented off-target amplification. However, all 4 qPCR replicates across two membranes for this air sample showed similar Ct values. This detection could rather represent an asymptomatic or pre-symptomatic individual shedding measles virus in the waiting room at this facility who did not ultimately obtain a measles diagnosis. While measles is not typically associated with asymptomatic carriers, some studies have suggested that asymptomatic infections may be more common than appreciated, although it is unknown if asymptomatic infections correspond with viral shedding.^12,13^ Measles vaccine strain was not detected in this sample, suggesting wildtype and not vaccine strain shedding.

Measles N450 sequences confirmed qPCR detections in three ED1 samples, including two of five from this site with Ct values below the qPCR limit of detection cutoff, and one with a Ct value higher than the cutoff. Thus, the limit of detection cutoff may have excluded some true positive samples. An assay with higher specificity for environmental samples may yield more sensitive results. Sequences were not produced from the ED2 detection sample despite multiple attempts, however, N450 sequencing was less sensitive than qPCR detection (3 qPCR positive samples from ED1 were unable to be sequenced), so N450 sequencing may not be reliably used for qPCR detection confirmation.

Measles sequencing from ED1 air samples yielded one sequence that was two nucleotides diverged from two other air sample sequences and all sequences derived from case patients during this outbreak, which were all identical. Measles virus was not able to be sequenced from five case patients from this outbreak, however, none of these patients were hospitalized at ED1. Thus, this diverging sequence could originate from a person who was shedding virus while at ED1, but was not tested for measles. Given the low N450 sequence diversity typically observed in measles outbreaks, this sequence could reflect a non-outbreak-associated person shedding measles. However, two nucleotide differences does not exclude the possibility of this sequence originating from an outbreak-associated infection. Together, the qPCR detection in ED2 and diverging N450 sequence detection indicate that air sampling may have detected additional measles cases that were not detected by passive measles surveillance or active outbreak case surveillance. Ultimately, the source and epidemiological significance of these detections could not be determined.

This study had several limitations. First, air sampler placements and weekly sampling durations were selected for other surveillance activities and were not optimized for this study. Shorter-term sampling and placement near measles patient rooms or in hallways used for transport may have been more informative. Second, nucleic acid detections used here do not indicate whether measles detected in the air represent intact or infectious virus. Specialized air samplers are required for recovering replication competent virus and could be used when assessing transmission risk. Importantly, no measles secondary transmissions were identified at any hospital included in this study.

The ability to detect measles virus in the air of indoor spaces with measles cases has implications for the use of air sampling for measles surveillance and risk assessment. The definition of measles virus exposure is generally broad and sensitive, leading to extensive exposure assessments and follow-ups that burden hospital and public health resources during outbreaks.^4,14,15^ Air sampling may be useful for investigating exposure and risk assessment to inform more efficient responses. Further, air sampling may be useful as a supplemental surveillance approach in high-risk settings by providing an early signal of measles virus shedding, including potentially during the presymptomatic stage, that could prompt additional investigation or public health response.

## Data Availability

All data produced in the present work are contained in the manuscript.

## ACKNOWLEDGEMENTS

We gratefully acknowledge Shelby Daniel-Wayman and Stephanie Gretsch, Chicago Department of Public Health, for case data. We thank the Laboratory-Based Surveillance team at Chicago Department of Public Health for air surveillance program and specimen coordination, including Samantha Smith, Dorothy Wright, and Haifa Wahbeh. We thank members of the Regional Innovative Public Health Laboratory and Microbiome and Genomics Core Facility at Rush University Medical Center for processing of air samples and generation of qPCR and sequencing data, especially Stefan Green, Giancarlo Balangue, Kevin Kunstman, and Jeremy Kahsen.

